# Stress, coping, and quality of life in the United States during the COVID-19 pandemic

**DOI:** 10.1101/2022.11.03.22281899

**Authors:** Fathima Wakeel, Jacelyn Hannah, Leah Gorfinkel

## Abstract

While research has widely explored stress, coping, and quality of life (QOL) individually and the potential links between them, there is a critical dearth in the literature regarding these constructs in the context of the COVID-19 pandemic. Our study aims to identify the salient stressors experienced, describe the coping strategies used, and examine the relationships between stress, coping, and current QOL among individuals during the pandemic. Data are from a nationally representative sample of 1,004 respondents who completed an online survey. Key measures included stressful life events (SLEs), coping strategies, and the physical and psychological health domains of QOL. Staged multivariate linear regression analyses examined the relationships between the two QOL domains and SLEs, controlling for sociodemographic and pre-existing health conditions and testing for the effects of coping strategies on these relationships. The most common SLEs experienced during the pandemic were a decrease in financial status, personal injury or illness, and change in living conditions. Problem-focused coping and emotion-focused coping were significantly related to higher levels of QOL, whereas avoidant coping was associated with lower QOL. Avoidant coping partially mediated the relationship between experiencing SLEs and reduced physical and psychological QOL. Our study informs clinical interventions to help individuals adopt healthy behaviors to effectively manage stressors, especially large-scale traumatic events like the pandemic. Our findings also call for public health and clinical interventions to address the long-term impacts of the most prevalent stressors experienced during the pandemic among vulnerable groups.

## Introduction

The COVID-19 pandemic has led to a prolonged period of stress due to its extensive adverse impacts, including increased mortality and morbidity, substantial economic challenges, heightened levels of uncertainty, and social isolation. Exposure to chronic stressors can significantly impact one’s health directly through the neuroendocrine system (i.e., recurring activation of neuroendocrine responses and consequent increases in cholesterol, blood sugar, triglycerides, and blood pressure) or immune system (i.e., impairment of the immune system and resulting risk of infection) pathways, as well as indirectly through unhealthy behaviors such as poor diet, smoking, substance misuse, and risky sexual behaviors [1–9]. Experiencing stressful life events (SLEs) has also been significantly associated with reduced quality of life (QOL) in a wide range of vulnerable populations, including racial and ethnic minorities [10], elderly persons [11,12], chronically ill patients [13], and children [14,15].

The World Health Organization [16] defines the quality of life (QOL) as “individuals’ perception of their position in life in the context of the culture and value systems in which they live, and in relation to their goals, expectations, standards and concerns” (p.1405). Some research has indicated that individuals have experienced decreased QOL during the pandemic. For example, stressors, such as loss of income, personal health effects, social isolation, and COVID-19 diagnosis, were negatively correlated with QOL during the pandemic [17]. Further, a study [18] in Germany found that the pandemic did not affect the QOL of individuals equally, with women, job seekers, and younger people reporting a significantly lower QOL. This study also indicated an overall decline in reported physical and psychological QOL during the pandemic [18].

Individuals cope with stressors in various ways. Three coping strategies widely investigated in the literature include problem-focused coping, emotion-focused focusing, and avoidant coping. Each of these strategies entails different methods for dealing with SLEs. Problem-focused coping involves stress-reducing tactics such as problem-solving, obtaining instrumental support, and planning [19,20]. Emotion-focused coping strategies include the use of emotional support, humor, religion, and positive reframing [19,20]. Avoidant coping involves behaviors such as substance use, distractions, and behavioral disengagement [21]. The literature has demonstrated that problem-focused coping is the most effective in stressful situations because it entails taking control of the stressor and using proactive methods to address it [20]. On the other hand, emotion-focused coping has been shown to be most effective when the stressor, such as a death of a loved one, is outside of one’s control [20,22]. Avoidant coping is the least effective and most harmful because it not only does not remove the stressor but also likely worsens existing stress, anxiety, and depression [23–25].

How one utilizes the different coping strategies can impact their QOL. Though several studies have captured how frequently various coping strategies were used during the COVID-19 pandemic, there is limited research on the relationships between coping strategies and QOL during this time period. A large-scale study [26] in the United Kingdom found that emotion-focused coping strategies were more likely to be used when individuals experienced financial stressors (i.e., their or their partner’s loss of employment or inability to work, decrease in household income) or worries about contracting or becoming severely sick from COVID-19 and that both problem-focused coping and avoidant coping strategies were likely to be employed when respondents reported adverse financial events as well as worries about finances, basic needs, or getting COVID-19. In terms of potential links between coping and QOL during the pandemic, research [27] has shown that for patients hospitalized with COVID-19, the use of problem-focused coping mechanisms had a direct and positive correlation with their QOL. Additionally, Quiroga-Garza et al. [28] found that problem-focused and emotion-focused coping were marginally, but significantly, correlated with well-being. Further, Shamblaw et al. [25] found that approach coping, which entails more proactive aspects of problem-focused and emotion-focused coping (e.g., planning, positive reframing, and use of emotional support), was associated with higher QOL, whereas avoidant coping was related to significantly reduced QOL during the pandemic.

While researchers have widely explored stress, coping, and QOL individually as well as potential links between them, there is a critical dearth in the literature regarding these constructs in the context of the COVID-19 pandemic. As the pandemic has had far-reaching and multidimensional health impacts on the population, it provides a unique opportunity to investigate how individuals cope with stressors during a relatively brief period of time and how different coping strategies may have differential impacts on individuals’ QOL. Our study uses a nationally representative sample of over 1000 U.S. adults to 1) Identify the salient stressors reported by individuals during the pandemic; 2) Describe the types of coping strategies used by individuals during the pandemic; and 3) Examine the relationships between stress, coping, and QOL among individuals during the pandemic.

## Materials and Methods

### Sample and Procedures

Data for this study are from a nationally representative sample of 1,004 respondents who completed our 25-30-minute online survey on *Prolific*, a web-based survey recruitment platform, in August 2021. *Prolific* creates nationally representative samples based on age, sex, and ethnicity data from the US Census Bureau. Survey questions were grouped into three main categories: 1) Well-being topics, including QOL, social relationship factors, and stress and coping; 2) Information topics, including sources of health information, telehealth services, and consumption behavior; and 3) Science and vaccines topics, including vaccine intentions and behaviors, attitudes and beliefs about scientific and medical research, and political and religious preferences. To reduce the potential respondent burden, respondents were randomly assigned two out of the three categories of questions. Therefore, though 1,500 respondents completed the overall survey, only 1,004 individuals completed the questions relevant to this study. This study was approved by the Lehigh University Institutional Review Board, and respondents provided informed consent before participating in the study.

### Measures

#### Quality of life

QOL was measured using the World Health Organization Quality of Life Abbreviated Version (WHOQOL-BREF) instrument [29]. This 26-item measure asks the respondent to reflect on various dimensions of their life, including general QOL, physical health, psychological health, social relationships, and environmental health, in the past four weeks using 5-point Likert scale responses. Scores for each QOL dimension were then rescaled to range from 0-100, per the instrument’s scoring instructions. In this analysis, we focused on the dimensions of physical health (7 items) and psychological health (6 items), as these dimensions measured perceptions regarding more internal experiences of health, as opposed to experiences with external factors such as social relationships and environmental situations. The physical and psychological health measures (α=0.82 for both) had good internal consistency (Table 1).

**Table 1:** Descriptive Statistics and Correlations between Study Measures.

|  | **Cronbach's alpha** | **Mean** | **SD** | **Range** | **1** | **2** | **3** | **4** | **5** | **6** |
| --- | --- | --- | --- | --- | --- | --- | --- | --- | --- | --- |
| 1. Stressful life events | N/A | 1.7 | 2.2 | 0-18 | 1.00 | 0.21* | 0.20* | 0.31* | -0.23* | -0.18* |
| 2. Problem-focused coping | 0.78 | 13.8 | 5.4 | 0-24 |  | 1.00 | 0.57* | 0.26* | 0.07* | 0.12* |
| 3. Emotion-focused coping | 0.67 | 11.1 | 5.1 | 0-24 |  |  | 1.00 | 0.33* | 0.03 | 0.14* |
| 4. Avoidant coping | 0.75 | 9.1 | 4.9 | 0-24 |  |  |  | 1.00 | -0.24* | -0.33* |
| 5. Quality of life (physical health) | 0.82 | 70.4 | 19.7 | 0-100 |  |  |  |  | 1.00 | 0.61* |
| 6. Quality of life (psychological health) | 0.82 | 64.1 | 21.3 | 0-100 |  |  |  |  |  | 1.00 |

\*p<0.05

#### Stress

Stress was measured using a modified version of the Holmes and Rahe Rating Scale [30], in which we asked respondents if they experienced any of the following 21 stressful life events (SLEs) during the pandemic: Death of spouse/partner, child, close family member, or close friend; personal injury or illness; domestic violence in the home; injury or illness in a child or other family member(s); divorce; separation from partner; imprisonment; loss of employment; loss of employment of spouse/partner; loss of educational opportunity; pregnancy; pregnancy of spouse/partner; childbirth; childbirth by spouse/partner; decrease in financial status; need to cut the size meals or skip meals because there wasn’t enough money for food; homelessness; foreclosure of mortgage or loan; eviction; change in living conditions; and increase in the frequency of arguments at home or work. Respondents were asked to mark all that applied, and we used the following categories of responses in our analyses: 0, 1, 2, 3, and 4+ SLEs. We also examined the individual effects of each SLE in our multivariate regression models. SLEs were not operationalized as a composite score because over half of the respondents reported not experiencing any SLEs during the pandemic, and the distribution of the composite measure was wide.

#### Coping

Coping style was measured using a 16-item version of Carver’s Brief COPE measure [21]. Dias et al. [31] grouped the Brief COPE items into three coping styles: problem-focused coping, emotion-focused coping, and avoidant coping. In our survey, individuals were asked to indicate, using 5-point Likert scale responses, how they coped with stressors over the past year. Problem-focused coping included four items relating to active coping and the use of informational support. Emotion-focused coping included six items pertaining to emotional support, humor, and religion. Avoidant coping had six items relating to self-distraction, substance use, and behavioral disengagement. We rescaled the problem-focused coping scores to be out of 24 points to compare our findings more easily regarding different coping styles. The problem-focused, emotion-focused, and avoidant coping measures had acceptable internal consistencies with Cronbach alphas of 0.78, 0.67, and 0.75, respectively (Table 1).

#### Sociodemographic variables

The following sociodemographic variables were included in the analyses: race/ethnicity, gender identity, annual household income, age, and marital status.

Race/ethnicity categories included American Indian or Alaska Native; Asian; Black or African American; Hispanic, Latino or Spanish Origin; Middle Eastern or North African; Native Hawaiian or Other Pacific Islander; White; other; and prefer not to say. Due to their low frequencies in our sample, we categorized American Indian or Alaska Native, Middle Eastern or North African, Native Hawaiian or Other Pacific Islander, and Other as “Other Race/ethnicity” for the analyses.

We included the following gender identity categories in the survey: cisgender male; cisgender female; transgender male; transgender female; non-binary/gender non-conforming; do not identify as female, male, or transgender; and prefer not to say. Our analysis grouped transgender, non-binary/gender non-conforming, and non-identifying individuals as “other gender identity” due to their low frequencies in the sample.

Annual household income was operationalized as the following categories: Less than $25,000; $25,000-$34,999; $35,000-$49,999; $50,000-$74,999; $75,000-$99,999; $100,000-$149,999; $150,000-$199,999; greater than or equal to $200,000; and prefer not to say. The analysis grouped individuals with annual household incomes of $150,000 to $199,999 and $200,000 or more due to their smaller frequencies.

Age was a continuous variable that was provided as an open-ended response.

Marital status categories included: single/never married; married; not married, but in a relationship and living with your partner; not married, but in a relationship and not living with your partner; separated; divorced; widowed; and other.

#### Pre-existing health conditions

Pre-existing health conditions that were included in the analyses were having a chronic physical condition, mental health condition, or disability.

Chronic physical health condition was operationalized as a dichotomous variable (i.e., any versus none) indicating whether the respondent reported being diagnosed with at least one of the following chronic illnesses: Multiple sclerosis; high blood pressure; COPD; diabetes; heart disease (heart failure, aFib, etc.); cancer; autoimmune (Psoriatic disease, Crohn’s/Ulcerative Colitis, etc.); asthma; rheumatoid arthritis; and other.

Mental health condition was a dichotomous variable (any versus none); conditions included the following: Mood disorder (e.g., depression, bipolar disorder, etc.); anxiety disorder (e.g., obsessive-compulsive disorder, panic disorder, phobias, etc.); eating disorder (e.g., anorexia, bulimia, etc.); Post-traumatic Stress Disorder; and other.

Disability was operationalized as a dichotomous variable (any versus none) indicating if the respondent reported being diagnosed with any of the following disabilities: Sensory impairment (vision or hearing); mobility impairment; learning disability (e.g., ADHD, dyslexia); and other.

### Analytical Approach

The analyses were conducted using RStudio, version 4.1.2 [32]. Frequency distributions of SLEs were obtained (Figure 1). The internal consistency (i.e., Cronbach’s alphas) of the study measures, as well as correlations (i.e., Pearson product-moment correlation coefficients) between the measures, were calculated (Table 1). Kruskal-Wallis tests were conducted to compare the means of QOL, SLEs, and coping strategy measures by sociodemographic and health-related characteristics (Table 2). Staged multivariate linear regression analyses were then conducted to examine the relationships between each of the two QOL dimensions (i.e., psychological health and physical health) and SLEs, controlling for sociodemographic and pre-existing health conditions and testing for the effects of coping strategies on these relationships. The following covariates were included in the six models: 1) Model 1: SLE categories (i.e., 4+ events, three events, two events, one event); 2) Model 2: Model 1 covariates and sociodemographic variables; 3) Model 3: Model 2 covariates and pre-existing health conditions; 4) Model 4: Model 3 covariates and problem-focused coping; 5) Model 5: Model 4 covariates and emotion-focused coping; and 6) Model 6: Model 5 covariates and avoidant coping (Tables 3 and 4). Overall, the number of missing values (i.e., three in total) in the data were minimal and were imputed using the average of existing responses.

**Table 2:**
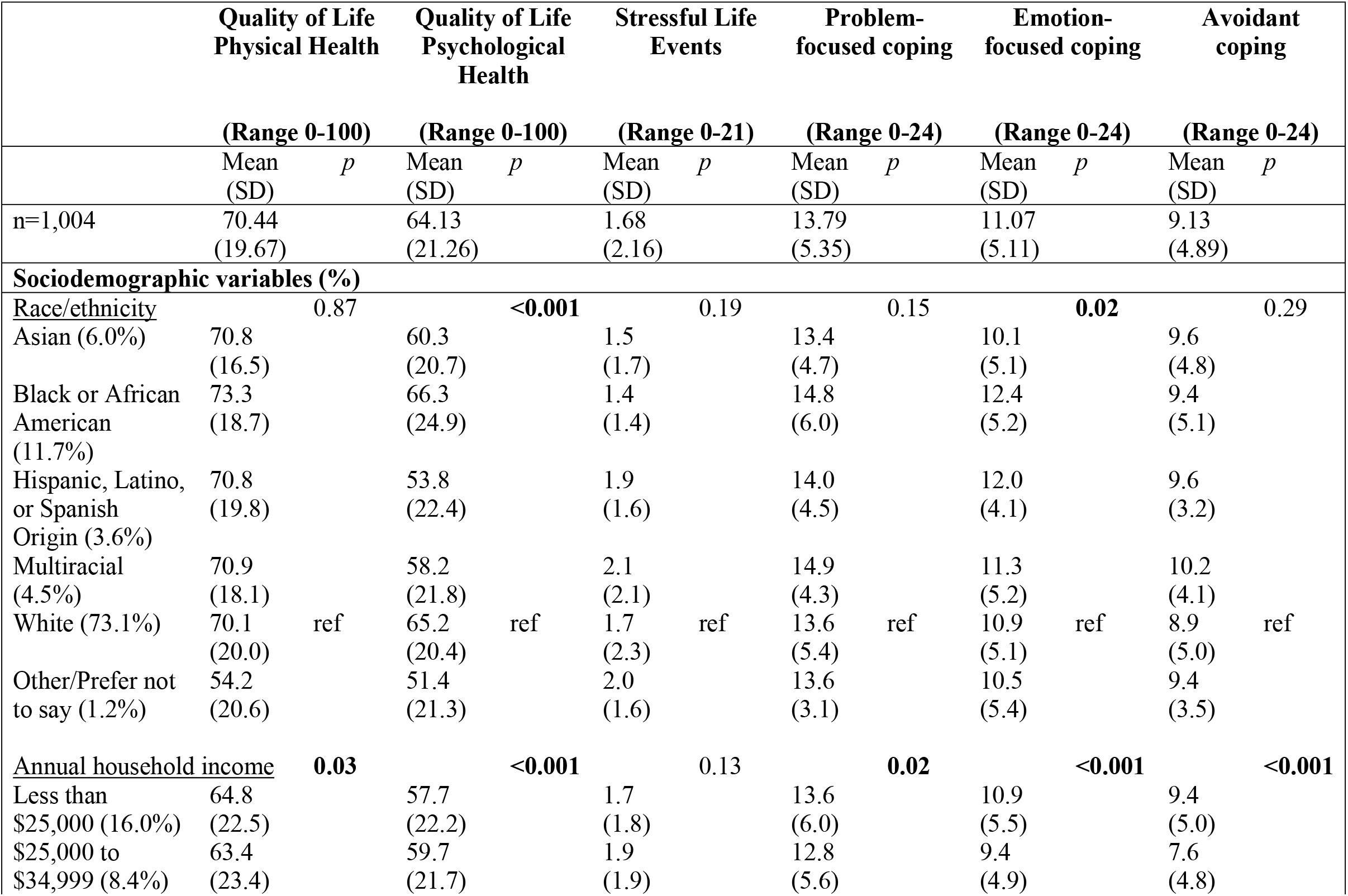
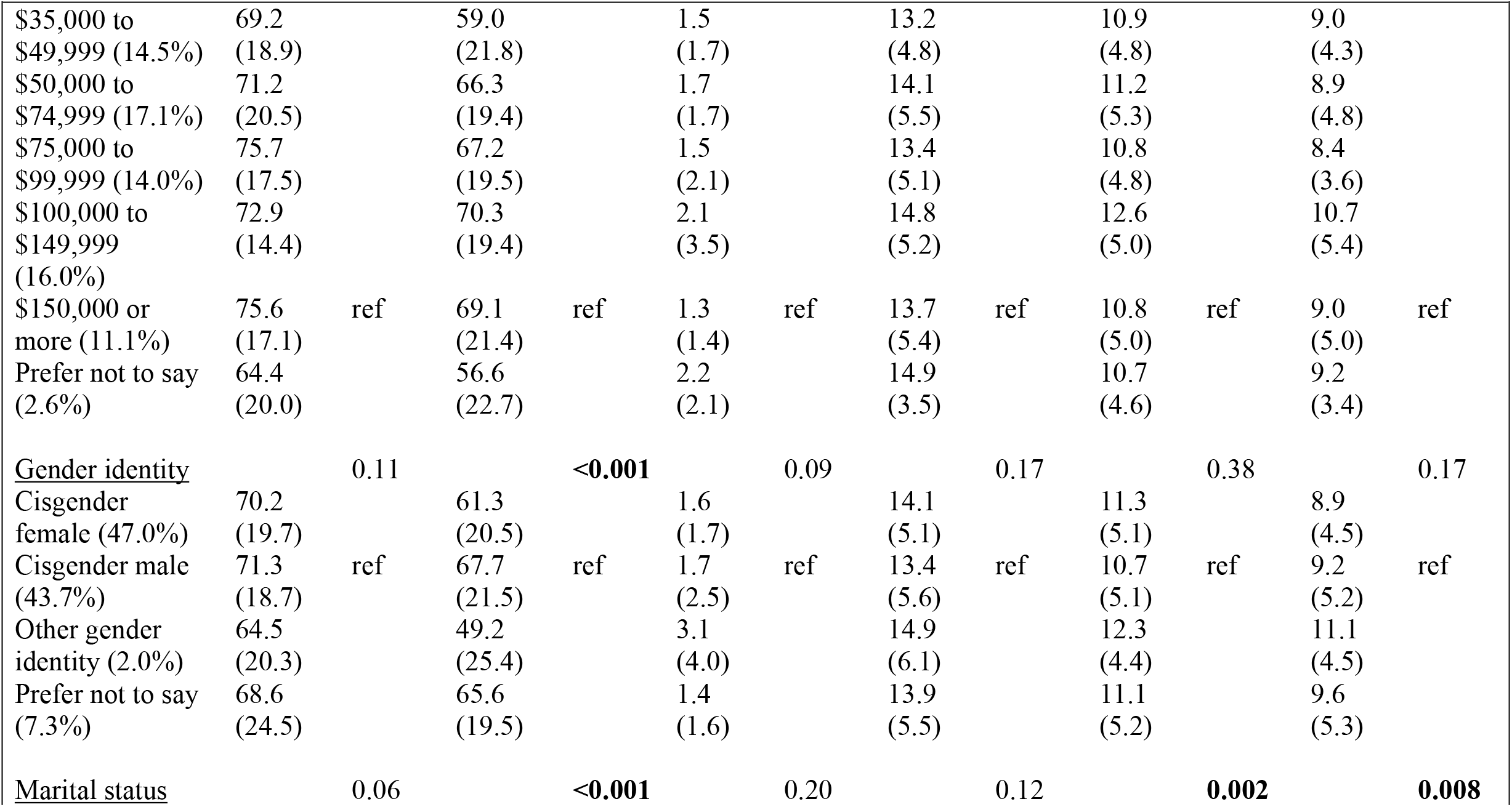
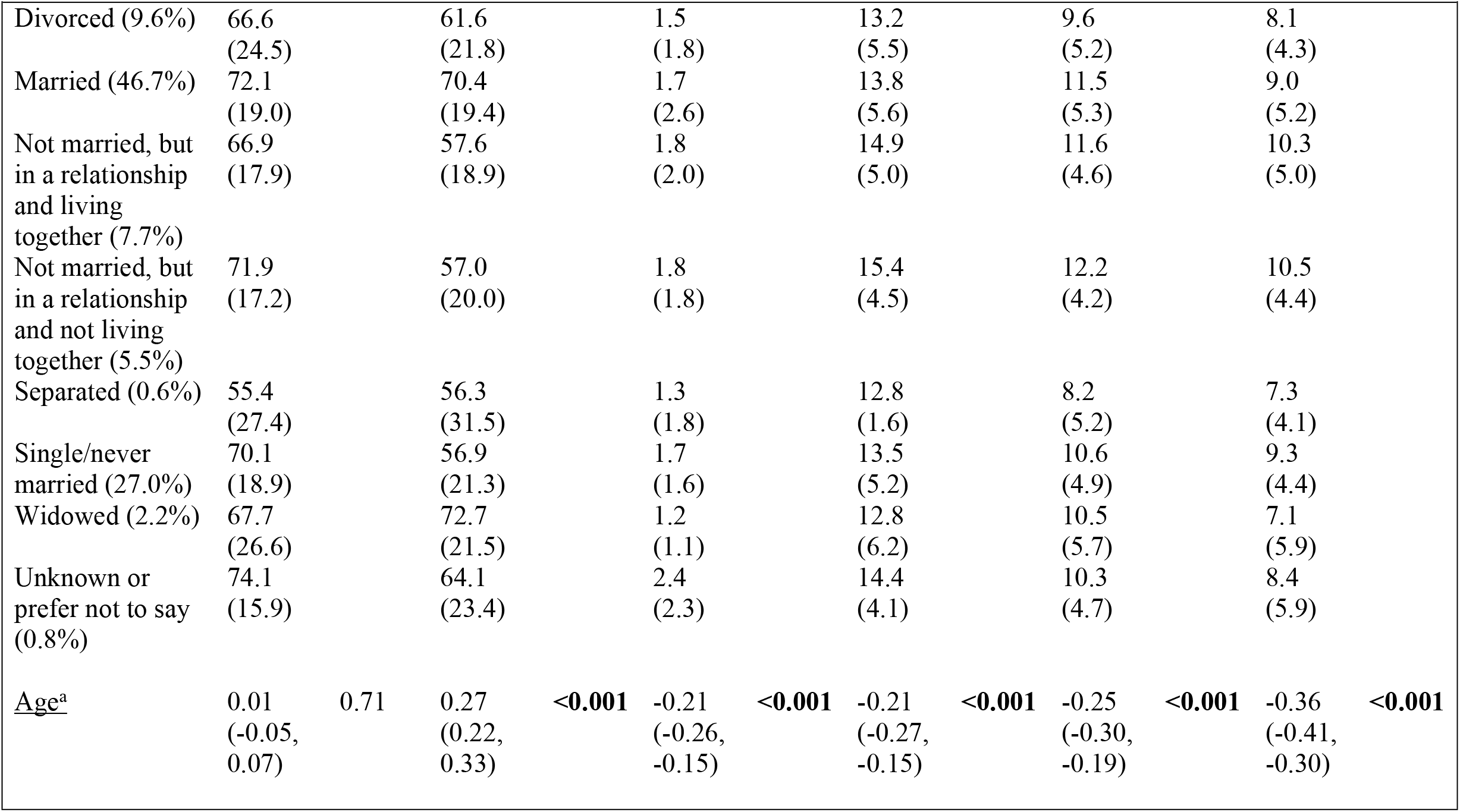
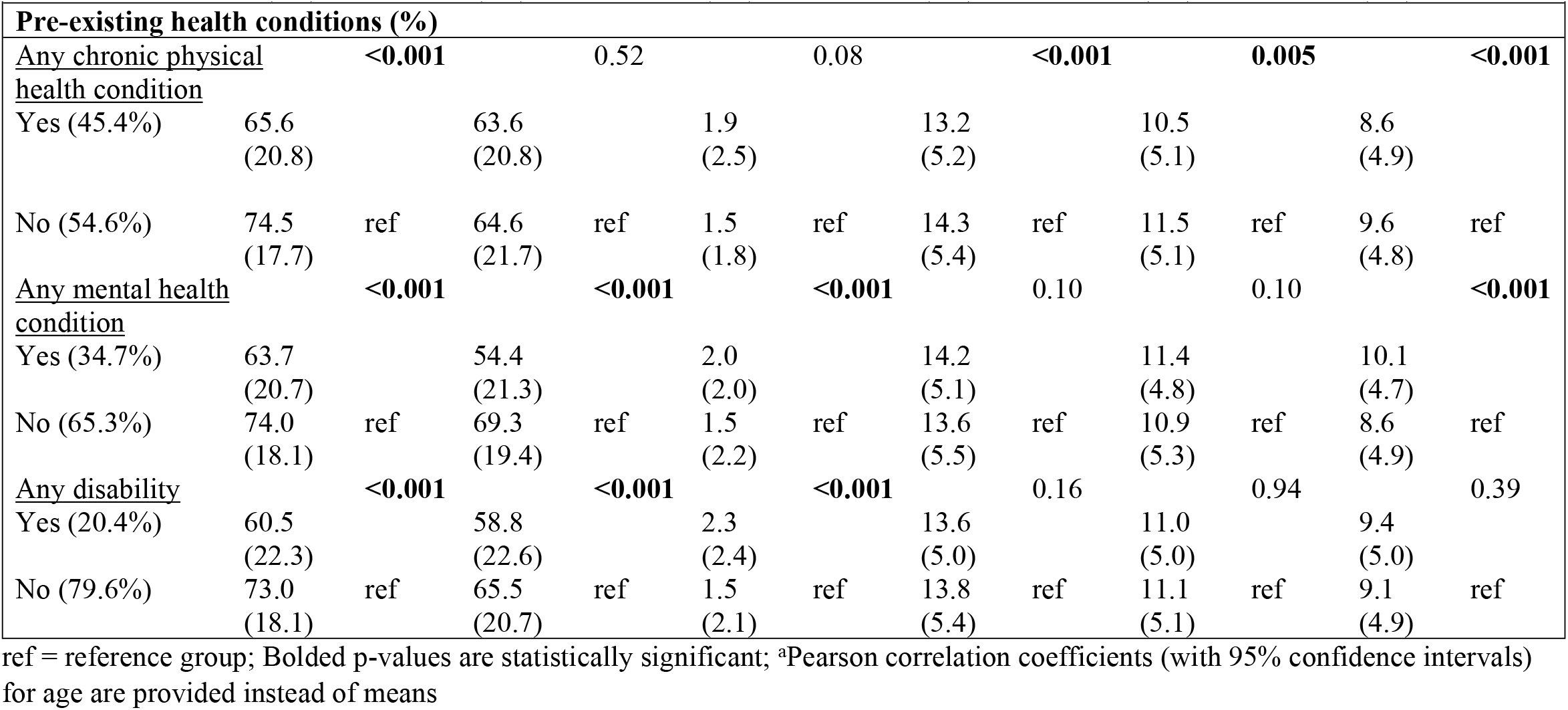
Means of Quality of Life, Stress, and Coping Strategies Scores by Sociodemographic and Health Characteristics.

**Table 3:**
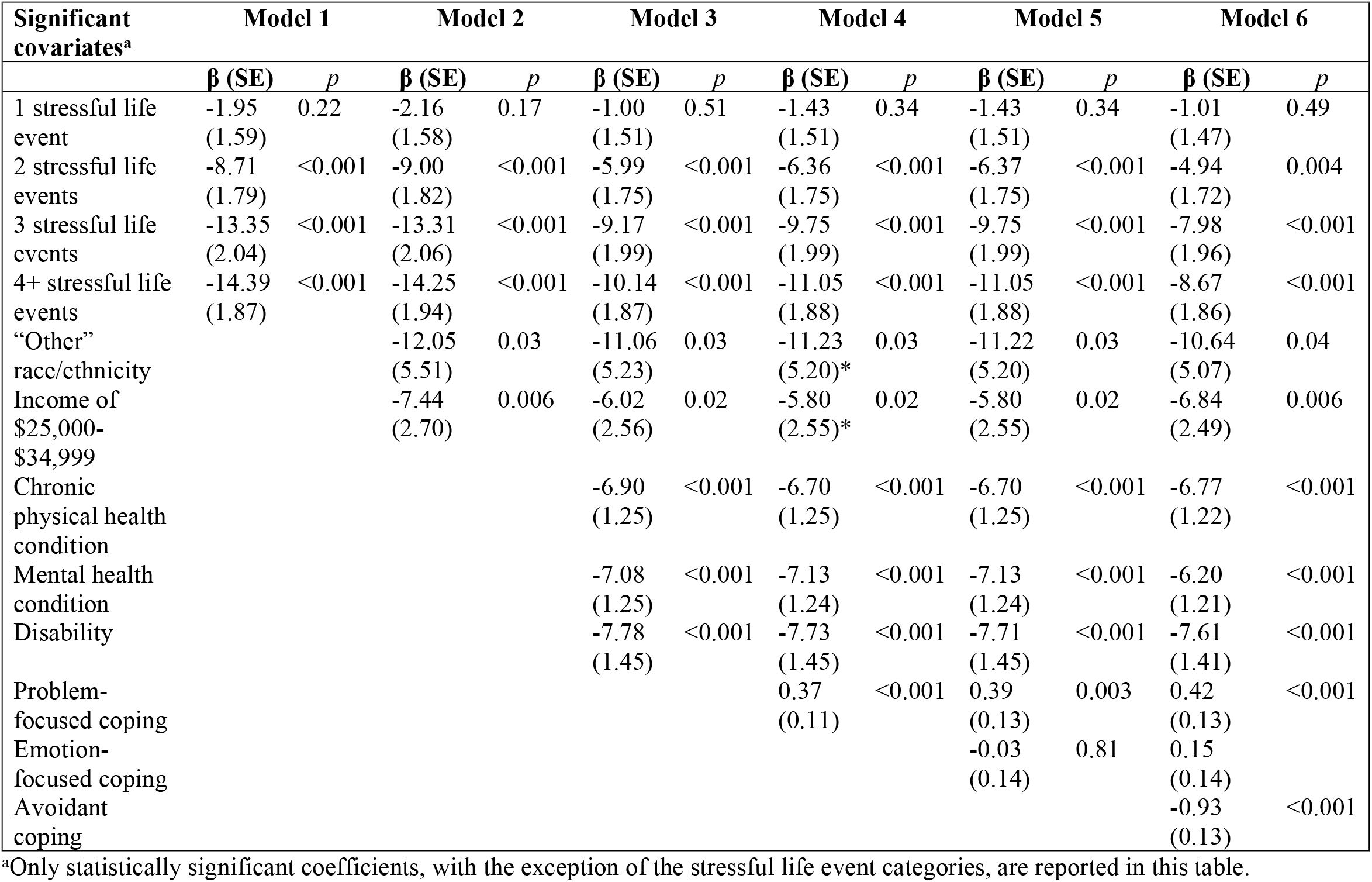
Relationships between Number of Stressful Life Events and Quality of Life (Physical Health)

**Table 4:**
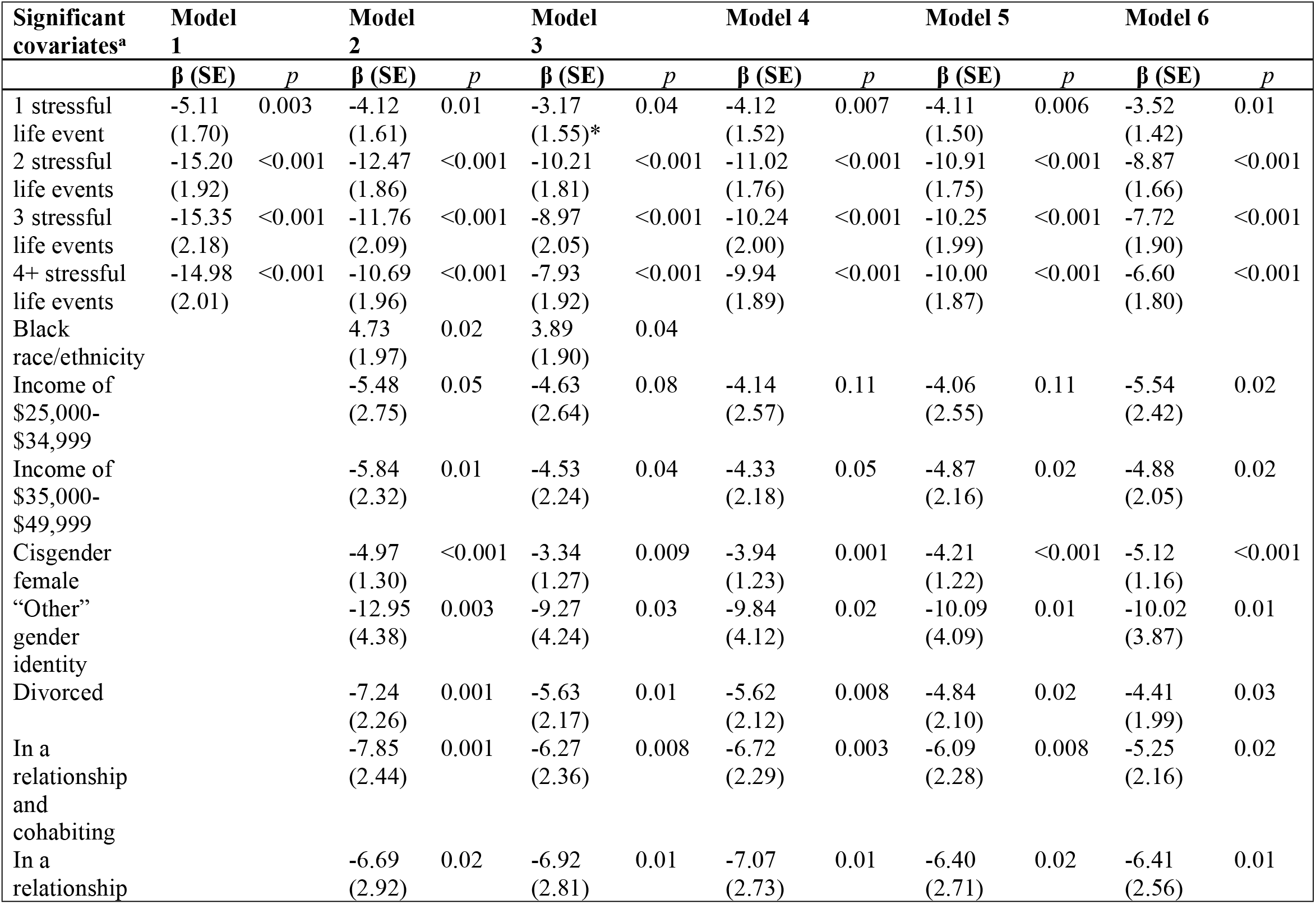
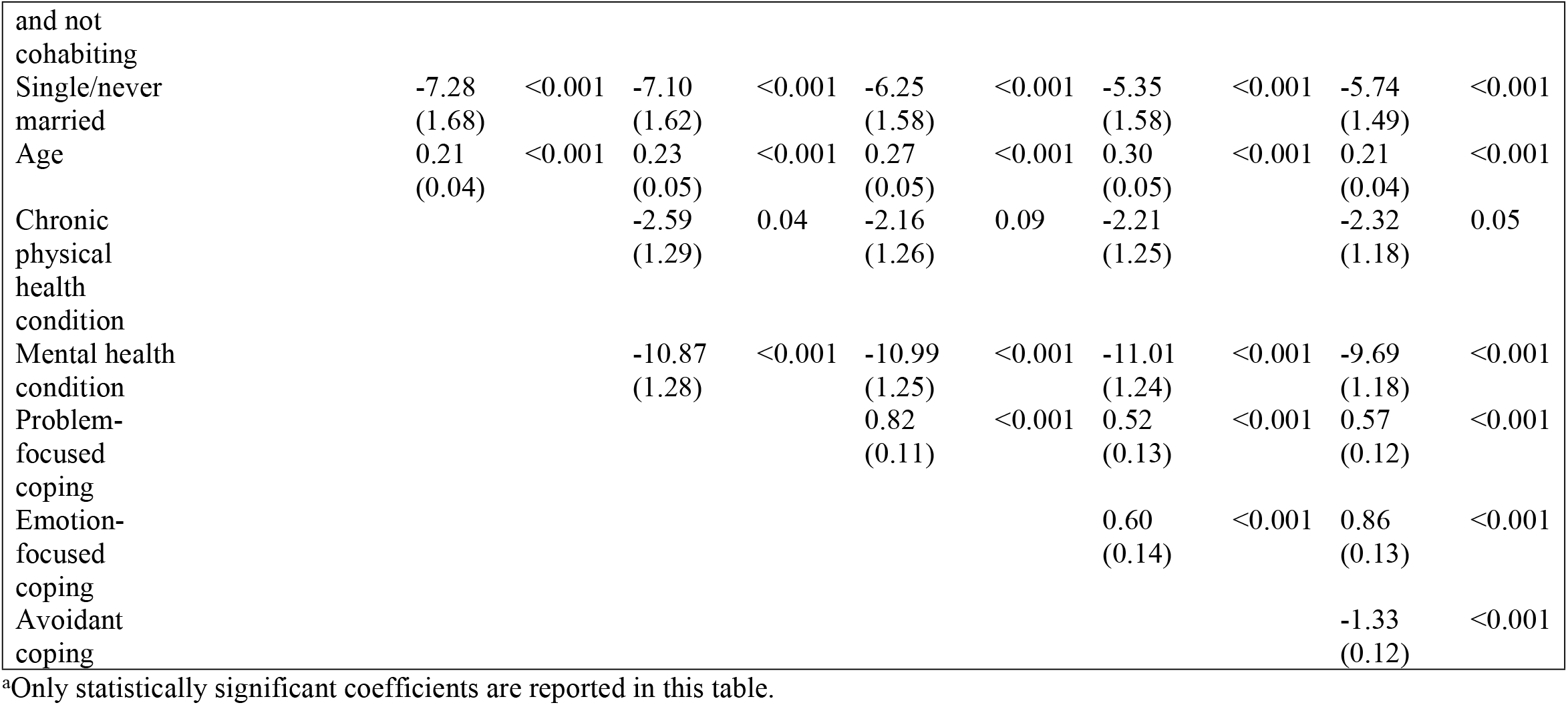
Relationships between Number of Stressful Life Events and Quality of Life (Psychological Health)

**Fig. 1:**
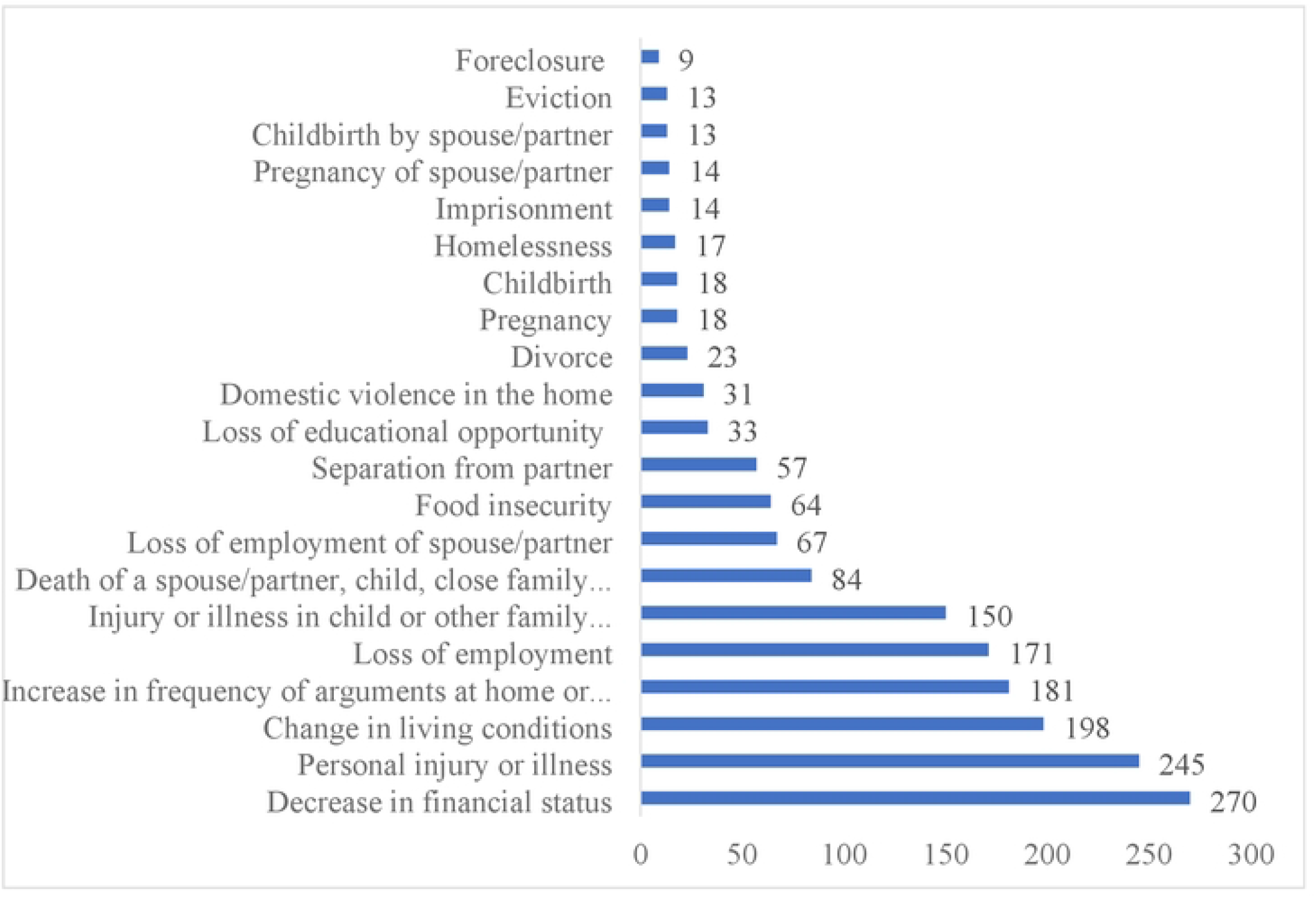
Frequency of Stressful Life Events during COVID-19 Pandemic. *Note:* Figure I demonstrates the frequency of stressful life events experienced by respondents during the COVID-19 pandemic

## Results

### Description of sample

The sample was predominantly White, followed by Black, Asian, Latinx, and multiracial individuals (Table 2). The sample was evenly spread out through the annual household income categories. The sample was relatively equally distributed by cisgender male and female, with only 2% of respondents in the “other” category (transgender, non-binary, or does not identify as male, female, or transgender). Almost 50% of the sample was married, and 27% were single/never married. The age distribution of respondents ranged from 18 to 82 years old, and the mean age was 44. Further, 42% of the sample reported having a chronic physical health condition, 35% reported having a mental health condition, and 20% reported having a disability.

The mean count of SLEs was 1.6, with a large spread within the sample ranging from zero to eighteen events (Table 2). The three most prevalent SLEs reported in the sample were a decrease in financial status, followed by personal injury or illness and a change in living conditions (Figure 1). Further, on average, the respondents reported higher levels of problem-focused coping, followed by emotion-focused coping and avoidant coping (Table 2).

### Bivariate analysis

Table 1 illustrates the descriptive statistics of the study measures as well as the correlations between measures. Bivariate analyses indicate that SLEs were positively associated with all three coping strategies and negatively associated with both QOL dimensions. Problem-focused coping was positively correlated with both QOL dimensions, emotion-focused coping was positively associated with QOL psychological health, and avoidant coping was negatively correlated with both QOL dimensions.

Significant unadjusted racial and ethnic differences in QOL psychological health and emotion-focused coping were found (Table 2). Latinx individuals reported the lowest QOL psychological health, whereas Asian individuals reported the lowest emotion-focused coping scores. Black individuals reported the highest scores for both measures. Further, there were significant differences by annual household income in QOL physical health, QOL psychological health, and all three coping strategies. Reported QOL generally increased with income levels. No consistently identifiable pattern emerged with coping strategies; individuals who had an income between $25,000 and $35,000 reported the lowest scores for all three types of coping, and individuals who had an income of $100,000-$150,000 generally reported the highest scores for all three types of coping. Additionally, the only significant differences in the gender identity variable were observed in QOL psychological health, with those who identified as “other” gender identity reporting the lowest QOL and cisgender males reporting the highest QOL.

There were significant differences by marital status for QOL psychological health, emotion-focused coping, and avoidant coping. Separated individuals generally reported the lowest scores across these measures, and those in a relationship but not cohabiting reported the highest emotion-focused and avoidant coping scores. Married and widowed individuals reported the highest QOL psychological health (Table 2). Further, those with a chronic health condition, mental health condition, or disability reported significantly lower QOL physical health. Individuals with a mental health condition or disability also reported significantly lower QOL psychological health and a higher number of SLEs. Those with a chronic health condition reported significantly lower scores for the three types of coping, and those with a mental health condition reported higher avoidant coping scores.

### Multivariate analysis

Staged multivariate linear regression analyses examined the relationships between QOL (the physical and psychological health dimensions), SLEs, and the three coping strategies. Table 3 displays the statistically significant covariates in each of the models for which the QOL physical health dimension was the outcome variable. Based on the final model (model 6), experiencing a higher number of SLEs was associated with increasingly reduced QOL physical health. After adjusting for all other covariates, problem-focused coping was positively associated with QOL physical health, whereas avoidant coping was negatively associated with this outcome. Other significant covariates that were associated with significantly reduced QOL physical health included “other” race/ethnicity, having an annual household income between $25,000-$34,999, and having a chronic physical health condition, mental health condition, or disability. Further, the final model revealed that avoidant coping is a potential partial mediator in the relationships between the QOL physical health dimension and experiencing 2, 3, and 4+ SLEs, with reductions in the SLE coefficients with the inclusion of avoidant coping. Figure 2A illustrates the partial mediation model for the relationship between experiencing 4+ SLEs, avoidant coping, and QOL physical health. Sensitivity analysis through Sobel tests confirmed that avoidant coping partially mediated the relationships between experiencing 2, 3, and 4+ SLEs and QOL physical health.

**Fig. 2:**
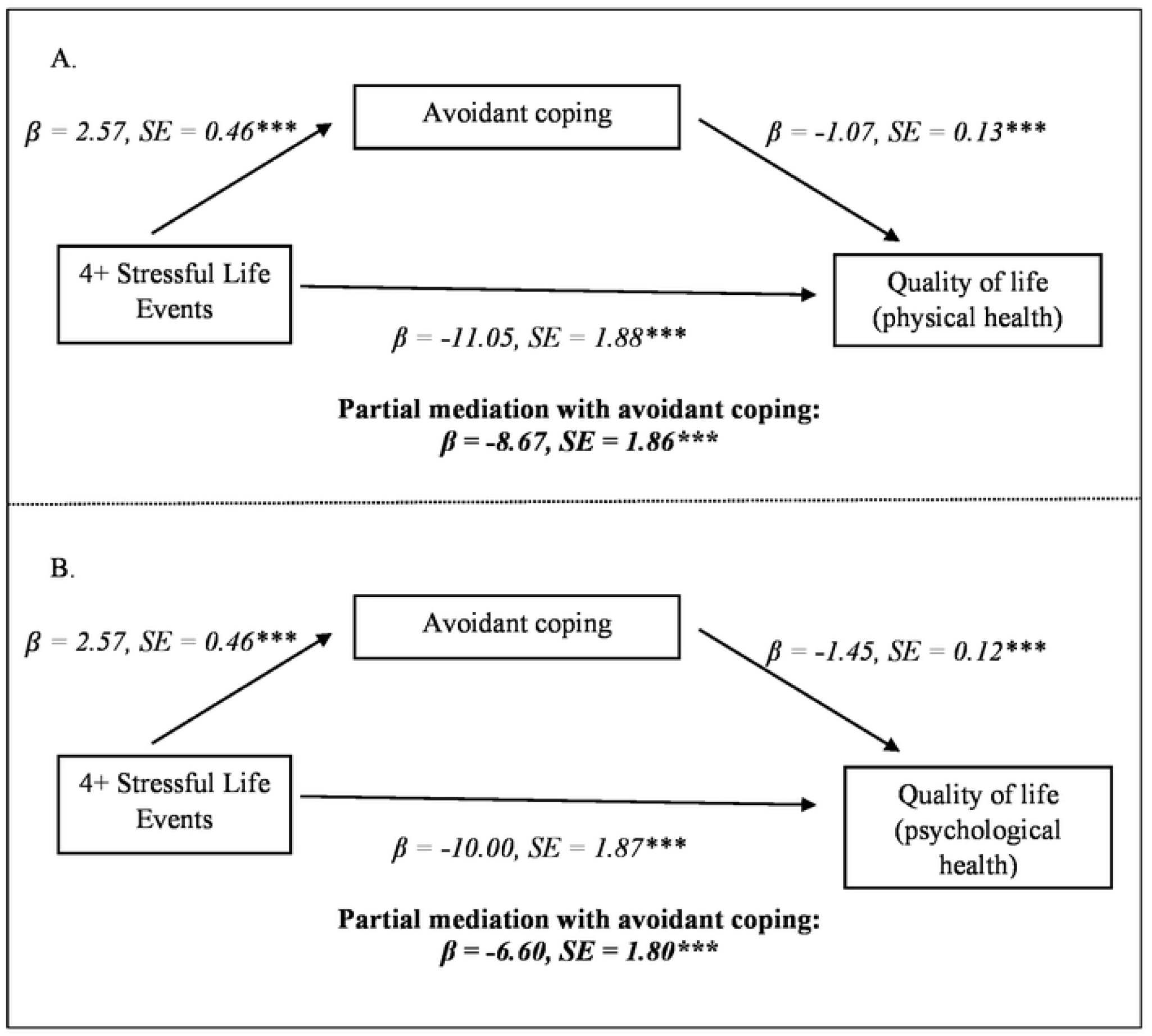
Mediation Models Relating Stressful Life Events to Quality of Life. *Note:* Figure 2A demonstrates that avoidant coping partially mediates the relationship between experiencing 4+ stressful life events and quality of life (physical health) during the pandemic. Figure 2B shows that avoidant coping partially mediates the relationship between experiencing 4+ stressful life events and quality of life (psychological health) during the pandemic. Standardized coefficients and standard errors are shown. *p<0.05; **p<0.01; ***p<0.001

Table 4 shows the staged linear regression models for the relationships between SLEs and the QOL psychological health dimension. In the final model, both problem-focused coping and emotion-focused coping were positively associated with QOL psychological health, while avoidant coping was negatively associated with QOL psychological health. Individuals who had an annual household income between $25,000-$49,999, those who were cisgender female or had an “other” gender identity, those who were divorced, unmarried but in a relationship, or single/never married, and those who had a chronic physical health condition or mental health condition had significantly lower QOL psychological health. On the other hand, increasing age had a small, positive association with QOL psychological health. Further, in the final model, experiencing 2 SLEs predicted the highest reduction in QOL psychological health, followed by experiencing 3 SLEs, 4+ SLEs, and 1 SLE. Again, avoidant coping partially mediated these relationships, with reductions in the SLE coefficients. The partial mediation model for the relationship between experiencing 4+ SLEs, avoidant coping, and QOL psychological health is shown in Figure 2B. Sobel test findings also indicated that avoidant coping partially mediated the relationships between experiencing 1, 2, 3, and 4+ SLEs and QOL psychological health.

## Discussion

This study contributes to the emerging literature on the psychological impacts of the COVID-19 pandemic by shedding light on individuals’ experiences of SLEs, utilization of various coping strategies, and current QOL during this traumatic global event. Our findings indicate that the most common SLEs experienced during the pandemic were a decrease in financial status, personal injury or illness, and change in living conditions. We also found that on average, respondents reported higher levels of problem-focused coping, followed by emotion-focused coping and avoidant coping. Further, problem-focused coping and emotion-focused coping were significantly related to higher levels of QOL, whereas avoidant coping was associated with lower QOL. Importantly, our study revealed that avoidant coping partially mediated the relationship between experiencing SLEs and reduced physical and psychological QOL.

Studies have reported conflicting findings regarding the most prevalent SLEs experienced by individuals during the pandemic. A U.S-based qualitative study by Jean-Baptiste et al. [33] found that the death of a loved one was the most common stressor experienced by respondents, followed by racism, discrimination including implicit bias and stereotyping, financial hardship and economic crisis, and personal health issues. On the other hand, a cross-sectional study conducted [34] in Iran found that the most prevalent stressor was the rise in essential good prices and that personal illness (i.e., being diagnosed with COVID-19) and the death of a loved one were ranked on the bottom of the list. Overall, our finding that experiencing financial difficulties and personal injury or illness were the most commonly experienced SLEs during the pandemic is consistent with this extant research.

Our findings corroborate existing literature indicating the positive association between the use of problem-focused coping and QOL pre-pandemic [35–37] and during the pandemic [25,27,28]. On the other hand, previous research, most of which was conducted pre-pandemic, has demonstrated inconsistent findings regarding the relationship between emotion-focused coping and QOL, with many studies pointing to a negative association between these two constructs [35,37,38]. A study conducted during the pandemic by Shamlaw et al. [25], however, has suggested that some emotion-focused coping strategies, such as the use of emotional support, may be related to well-being. As the pandemic instigated or exacerbated a wide range of unexpected and unpredictable stressors, such as personal illness, illness and deaths of loved ones, and unemployment, we posit that the use of emotion-focused coping was likely helpful in navigating these situations. Further, our findings regarding the inverse link between the use of avoidant coping strategies and QOL is supported by most extant literature [25,35,39,40]. Importantly, our study found that avoidant coping mediated the relationship between SLEs and reduced QOL during the pandemic. Some previous research has revealed similar findings in other contexts. For example, a study by Langford et al. [41] found that the avoidant coping strategy of disengagement coping mediated the association between SLEs and cancer-related distress.

Our study’s findings have important clinical and public health implications. Greater exposure to stressors was linked with avoidant coping strategies, which were, in turn, associated with significantly reduced QOL. Therefore, it is essential that mental healthcare and primary care providers dissuade the use of avoidant coping among patients, particularly those who experience elevated levels of stress. Alternatively, clinicians should promote the use of problem-focused coping, in general, to decrease the severity of stressors and the use of emotion-focused coping strategies when addressing uncontrollable or unpredictable stressors, such as large-scale traumatic events. Our study also highlights the most prevalent SLEs experienced during the pandemic. Hence, these findings call for public health and clinical interventions to address the long-term impacts of these stressors post-pandemic, especially among vulnerable groups such as racial/ethnic and gender minorities, cisgender women, lower-income individuals, unmarried individuals, and those with a chronic physical health condition, mental health condition, or disability.

Our study has some limitations that should be considered when interpreting its findings. First, as the survey was conducted in August 2021 and inquired about events that occurred over the time period of 17 months (since the lockdown in March 2020), responses may have been vulnerable to recall bias. Second, as this study was cross-sectional, causality cannot be assumed in the relationships between SLEs, coping strategies, and QOL. However, though respondents were asked to report on SLEs experienced and coping strategies used during the pandemic, responses regarding QOL inquired about current (i.e., in the past month) perceptions and feelings. Third, the sample, though nationally representative in terms of age, sex, and ethnicity, was not representative in terms of sociodemographic factors such as education or race. We partially addressed this limitation by not including education in our analysis; instead, we used income, which was more representative of the US population, as a proxy for socioeconomic status.

Despite these potential limitations, our study has critical implications for future research directions. Longitudinal research is needed to explore temporal relationships between the previous experience of SLEs, subsequent coping strategies that are employed, and current QOL. Further, as previous research has suggested that social support may act as a moderator in the relationship between stress and QOL [42,43], researchers should consider the roles of social support and sense of community in the relationships between SLEs, coping strategies, and QOL, including the social relationships dimension of QOL, during global stressors such as the COVID-19 pandemic. Along these lines, traumatic events have been shown to lead not only to stress but also to posttraumatic growth [44]. Therefore, future research should explore the development of both posttraumatic stress and growth after the pandemic, as well as their relationships with coping strategies and QOL.

## Conclusion

Our study contributes to the literature by being the first, to our knowledge, to indicate that avoidant coping mediated the relationship between experiencing SLEs and reduced physical and psychological QOL during the pandemic. Along these lines, we found that problem-focused coping and emotion-focused coping during the pandemic were significantly related to higher levels of current QOL, whereas avoidant coping was associated with lower QOL. Further, the most common SLEs experienced during the pandemic were a decrease in financial status, personal injury or illness, and change in living conditions. Our findings inform clinical interventions to help individuals adopt healthy behaviors to effectively manage stressors, especially large-scale traumatic events like the pandemic. Our study also sheds light on the most prevalent SLEs experienced during the pandemic, therefore calling for public health and clinical interventions to address the long-term impacts of these stressors post-pandemic, especially among vulnerable groups.

## Data Availability

The data that support the findings of this study are openly available in the Open Science Framework at https://osf.io/htazm/?view_only=5f563f24cadb4527ae6dcc2fe068a6db.

## Acknowledgements

The authors would like to thank the Lehigh University COVID-19 Nationwide Study Team for their assistance with this study.

